# Characteristics and outcomes of hospitalized adults with COVID-19 in Nepal: a multicenter, prospective cohort study

**DOI:** 10.1101/2020.10.03.20206128

**Authors:** Ashok Chaudhary, Uday Narayan Singh, Pramod Paudel, Niresh Thapa, Kamal Khadka, Prameshwar Kumar Sah, Sher Bahadur Kamar, Jagadish Joshi, Kamar Hasan Ansari, Shree Ram Tiwari, Sarbesh Sharma, Sanjay Kumar Jaiswal, Ramesh Joshi, Samikchya Baskota, Arjun Prasad Tiwari, Hem Raj Pandey

## Abstract

**Introduction:** There is limited data on clinical course and outcomes of hospitalized adults with COVID-19 in Nepal. Thus, it is imperative to characterize the features of this disease in the domestic context.

**Methodology:** We identified all adult patients with laboratory-confirmed COVID-19 admitted to five different hospitals in Nepal from June 15 to July 15, 2020. We collected epidemiological, socio-cultural and clinicopathologic data, and stratified the patients based on their symptom status.

**Results:** The study included 220 patients with an overall median age of 31.5 (25-37) years, and 181 (82.3%) were males. 159 (72.3%) were asymptomatic, and 163 (74.1%) were imported cases. Of 217 patients with the available data, 110 (50.7%) reported their annual household income less than 2000 US dollars, and 122 (56.2%) practiced Pranayama (yogic rhythmic breathing techniques) regularly. Eight patients (3.6%) required supplemental oxygen and two patients (0.9%) died. None of the patients who practiced Pranayama regularly required supplemental oxygen. Compared to asymptomatic patients, symptomatic patients had greater proportion of females (31.1% vs. 12.6%, p=0.001), imported cases (85.2% vs. 69.8%, p=0.02), illiterates (26.8% vs. 12.1%, p=0.01), alcohol users (43.3% vs. 24.5%, p=0.01), patients feeling stigmatized by society (45.8% vs. 22.6%, p=0.001), and had higher platelet count (253× 10^9^/L vs. 185×10^9^/L, p=0.02).

**Conclusions:** Most cases were imported, asymptomatic young males, with very few deaths. Pranayama practice was associated with protection against severe COVID-19, but more data is needed to substantiate this. The association of platelets count with symptom status in the Nepalese population needs further exploration.

## Introduction

The clinical course of coronavirus disease 2019 (COVID-19) varies with geographical regions [1]. In addition, different ethnicities have been differently affected by this disease, independent of geographic regions [2]. South Asia, one of the most densely populated regions of the world where nearly a quarter of the humanity live, is facing extraordinary challenges in dealing with this pandemic [3]. COVID-19 has precipitated more than just a health crisis in this region [4]. Several studies have shown that South Asians are more adversely affected by this disease compared to Chinese and Whites [5].

While all the exact reasons behind these differences are not known, it is logical to think that each country needs to have its tailored strategy based on the data from their local context [6]. Thus, there is a compelling need for domestic evidence on COVID-19 to make informed decisions and optimize the national strategy. However, there are relatively few prospective cohort studies on COVID-19 from South Asia, and none from Nepal, a lower-middle-income South Asian nation of nearly 30 million people [7], landlocked by China on North, and India on East, West and South. The data on the clinical course of hospitalized COVID-19 adults in Nepal is limited, and we do not have robust scientific documentation of how this disease behaves here compared to the other countries.

In this first multicenter, prospective cohort study on hospitalized adults with COVID-19 in Nepal, we describe the epidemiological, sociocultural, clinical characteristics and outcomes of adults with this disease admitted to five different hospitals of Nepal.

## Methodology

### Study design and setting

This was a multicenter, observational, prospective cohort study done at five different government-designated COVID-19 hospitals of Nepal. The hospitals are: 1. Seti Provincial Hospital, Dhangadhi, Kailali 2. Hospital of Karnali Academy of Health Sciences, Jumla 3. Rapti Provincial Hospital, Tulsipur, Dang 4. Bharatpur Hospital, Bharatpur, Chitwan 5. Narayani Hospital, Birgunj, Parsa. These hospitals are located in five different provinces out of seven in Nepal. As of August 18, 2020, more than 85% of the total cases in Nepal were reported from these five provinces where these hospitals are located. All, except Rapti Provincial Hospital, are Level 2 COVID-19 hospitals as per the designation of government. Rapti Provincial Hospital is a Level 1 hospital. As of August 18, 2020, there were a total of 14, 12 and 3 government-designated Level 1, 2 and 3 hospitals in Nepal respectively [8].

### Study population

We prospectively identified all laboratory-confirmed COVID-19 adult patients (aged ≥ 18 years) admitted to these hospitals from June 15 to July 15, 2020. The diagnosis of COVID-19 was confirmed by Reverse Transcriptase-Polymerase Chain Reaction (RT-PCR) technique on nasopharyngeal and/or throat swabs. Each patient was followed up until one of the three outcomes: discharge from the hospital after improving. referral to higher-level hospital, or in-hospital death. On July 31, the last patient of our study was discharged. We did not calculate the sample size as we included all eligible patients during the time-window of this study.

We took oral informed consent from the patients. Nepal Health Research Council approved and provided ethical clearance to this study (Approval ID: 443/2020 P). This study was conducted in accordance with the principles of Declaration of Helsinki.

### Data collection

We used standardized case report forms which were developed based on ideas generated from the WHO Clinical Case Report Form. There were two modules in the form. The first module was completed within 24 hours of admission, and the second module was completed at discharge, referral, or death. We interviewed the patients and collected data from their medical records to obtain epidemiological, sociocultural, demographic, clinicopathologic data at presentation. The data on clinical course, management and outcomes were collected at the time of discharge, death or referral of the patient. We categorized the patients according to their symptom status, asymptomatic versus symptomatic.

The decision regarding diagnostic and therapeutic aspects of patient management was solely at the discretion of treating physician at the individual hospitals and was not influenced by this study.

### Statistical Analysis

We used counts (percentages) to describe categorical variables and medians with interquartile range (IQR) for continuous variables. Mann-Whitney U (Wilcoxon rank-sum) test was used to compare the differences in continuous data between the two groups. For categorical data, Pearson’s chi-square test or Fisher’s exact test was used. No imputation of the missing data was done. A two-tailed P value <0.05 was considered statistically significant. Statistical analyses were performed using IBM SPSS Statistics, version 25.0 (IBM Corp., Armonk, NY, USA).

## Results

This study included 220 adult inpatients with COVID-19 confirmed by RT-PCR, admitted to five different hospitals of Nepal. All patients were Nepalese.

Table 1 summarizes the socio-demographic and clinical characteristics of the patients stratified by symptom status, asymptomatic versus symptomatic. Overall, the median age was 31.5 years (IQR 25-37), and 82.3% (181/220) were males. 4.5% (10/220) were healthcare workers.74.1% (163/220) had recently returned from the foreign country with most of them (139/220) returning from India.

**Table 1.**
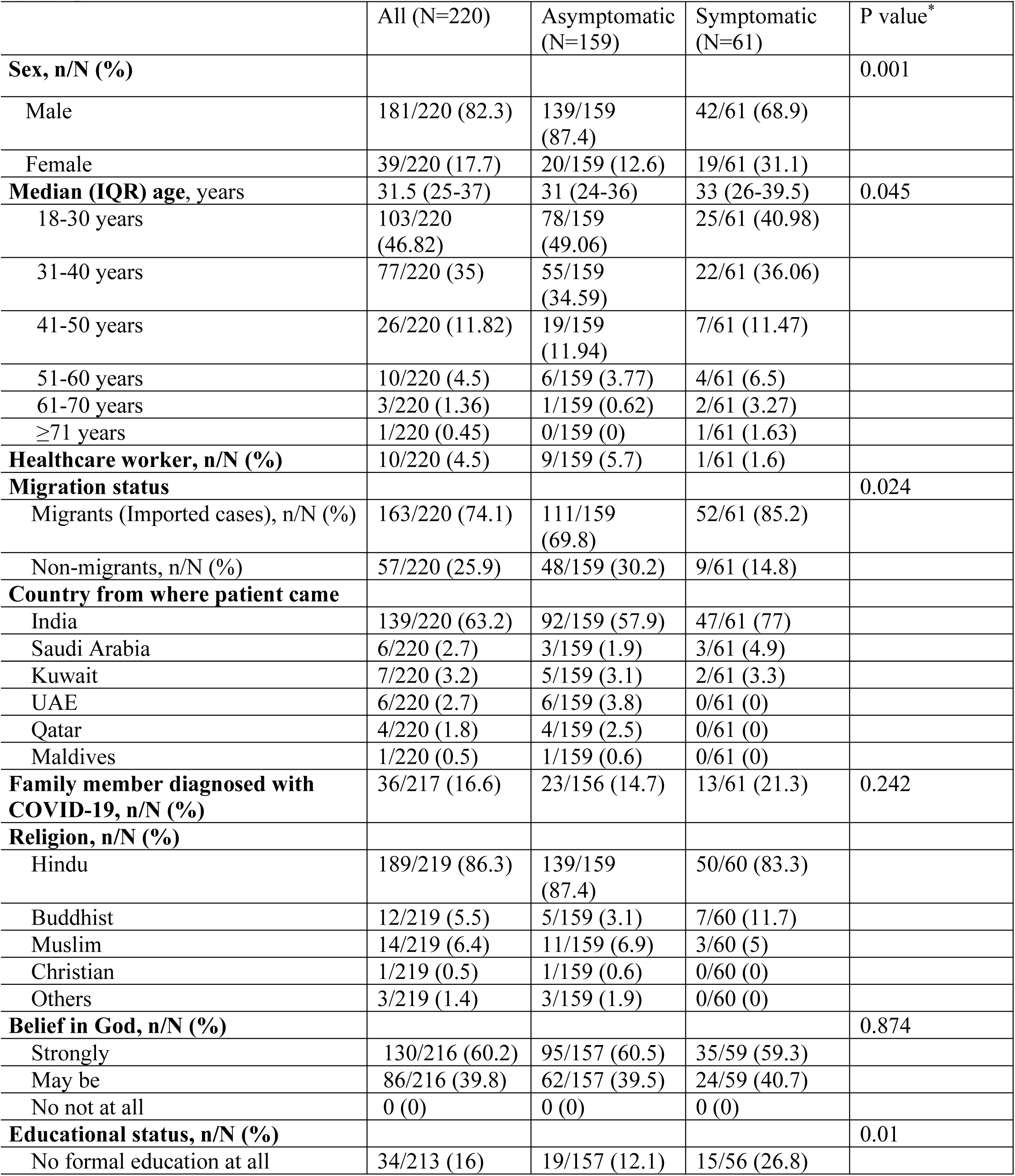
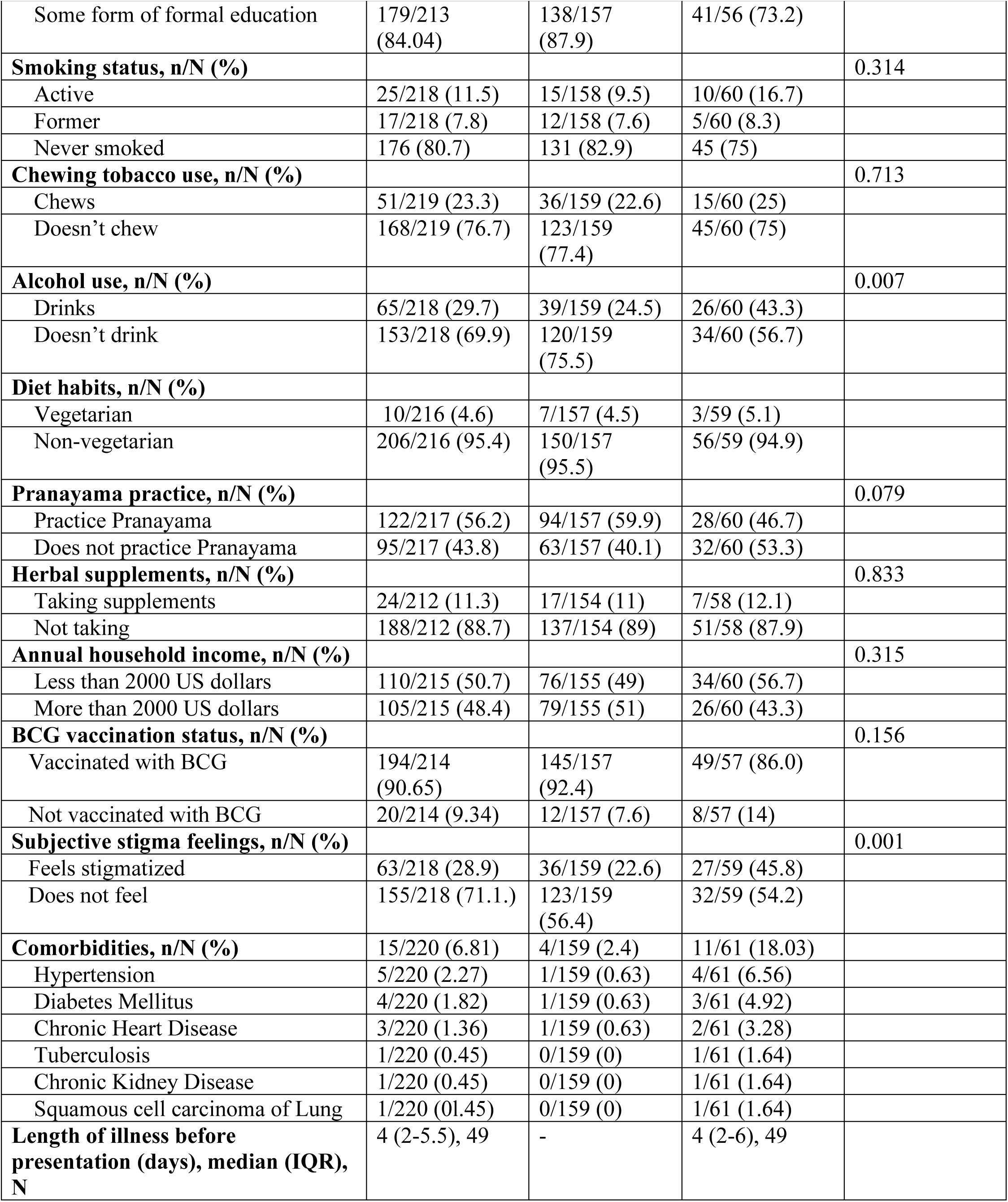
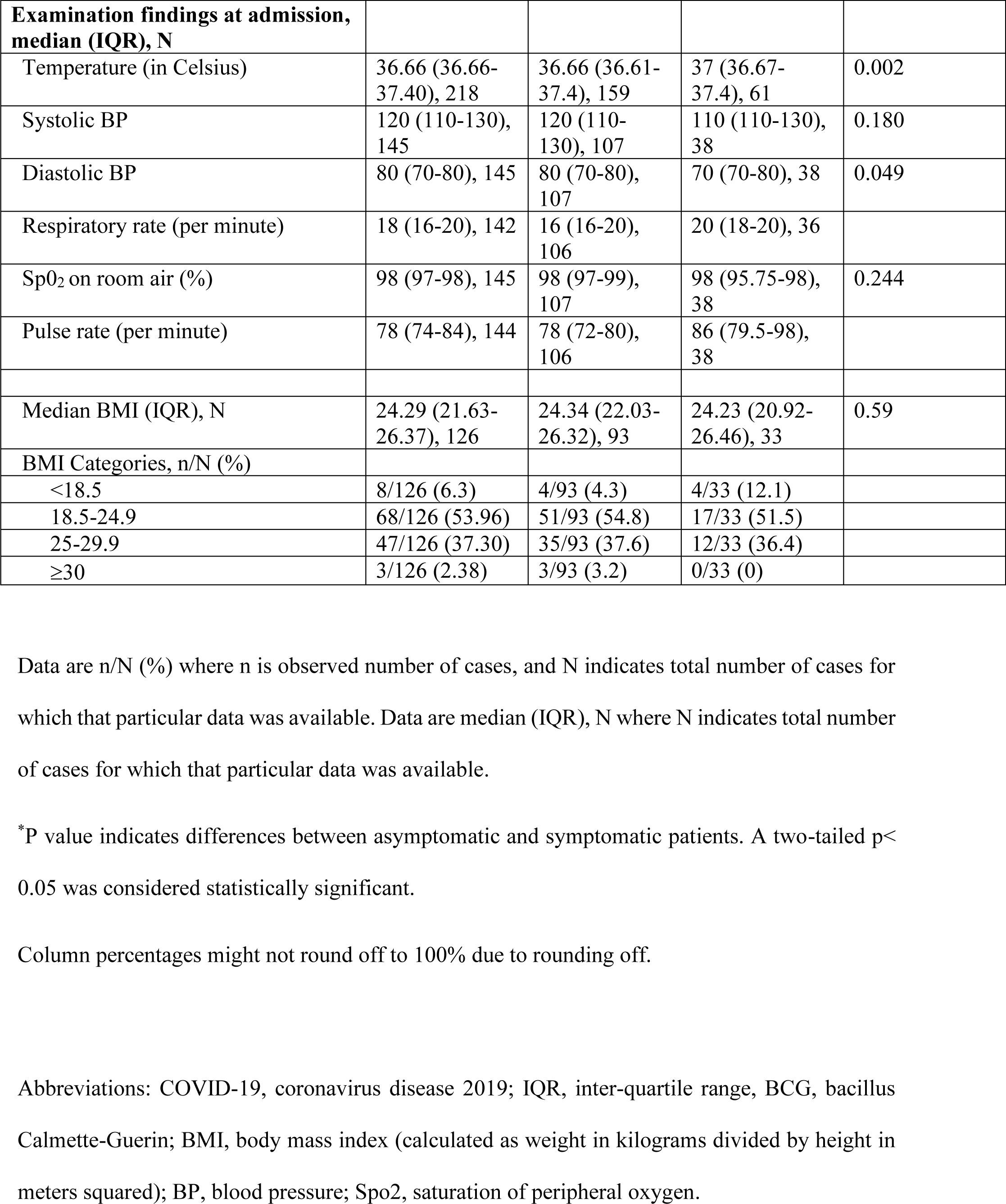
Characteristics of inpatients with COVID-19, admitted to five different hospitals of Nepal, stratified by symptom status.

Most of the patients were low skilled workers, with more than half (110/217) reporting their annual household income less than 2000 US dollars. One in six patients (36/217) had at least one family member diagnosed with COVID-19.86.3% (189/220) were Hindus. There were no non-believers of God.16% (34/213) had never been enrolled to any school.4.6% (10/216) were vegetarian by food habits. Of 214 patients with the available data, 90.7% (194/214) were vaccinated with Bacillus Calmette-Guerin (BCG) at the time of their birth. 56.2% (122/217) practised Pranayama regularly. Pranayama is a form of ancient yogic breathing technique and exercise. Just a little more than one in ten patients (24/212) took herbal supplements of any kind (table 1).

Of all patients, only 27.7% (61/220) were symptomatic at presentation. Figure 1 shows the frequency of symptoms. The median duration of symptoms before hospitalization was 4 days (IQR 2-5.5). 6.8% (15/220) had co-morbidities of some kind. Of 126 patients with the available data, the median BMI was 24.29 (IQR 21.6-26.4), with 6.3% (8/126) being underweight, and 2.38% (3/126) being obese.

**Figure 1.**
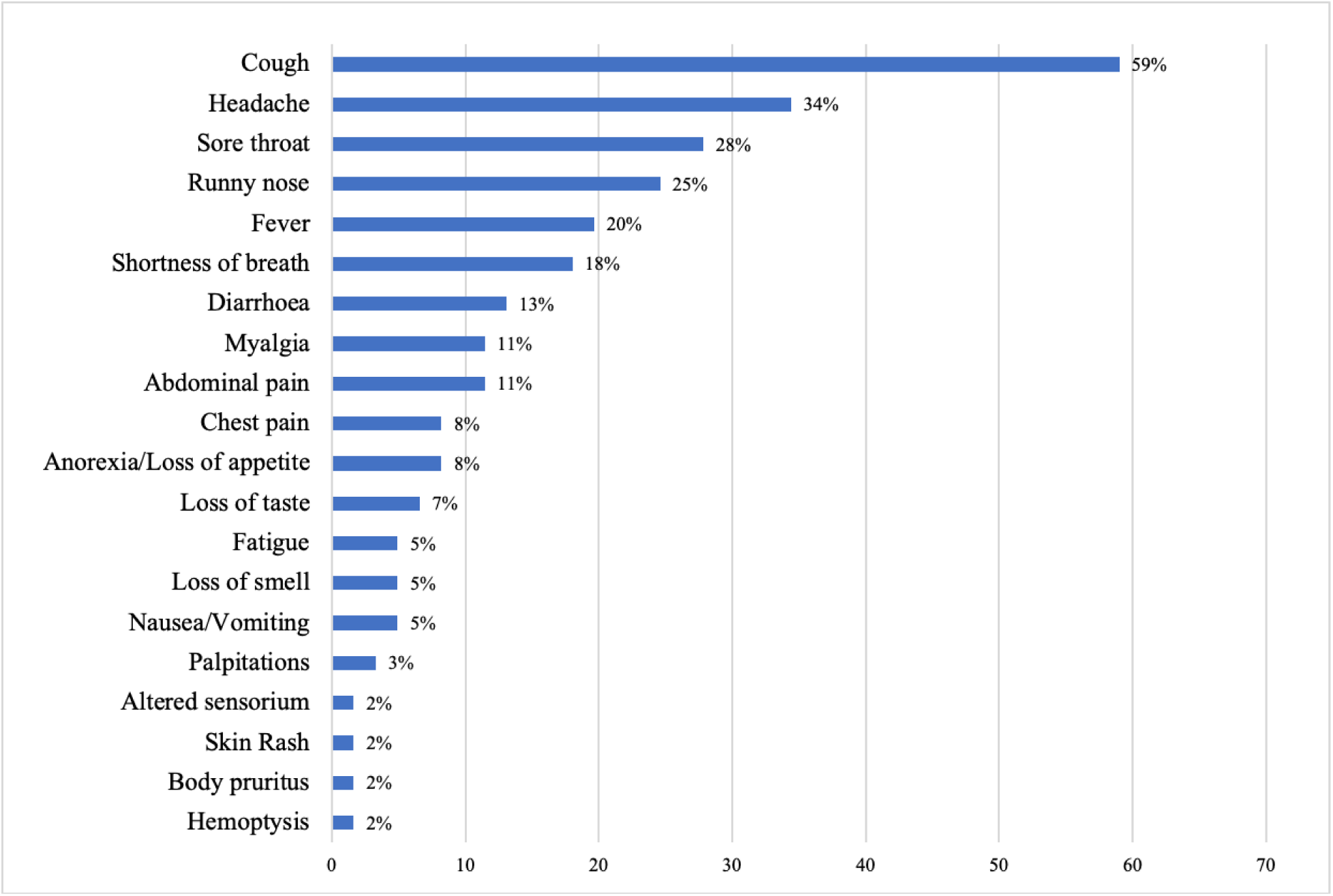
Symptoms of hospitalized adults COVID-19 patients in Nepal (N=61).

Compared to asymptomatic patients, symptomatic patients had higher proportion of females (31.1% vs. 12.6%, p=0.001), patients recently returning from abroad (85.2% vs. 69.8%, p=0.024), illiterate patients with no formal education at all (26.8% vs. 12.1%, p=0.01), patients who drink alcohol (43.3% vs. 24.5%, p=0.007), patients who felt stigmatized by society (45.8% vs. 22.6%, p=0.001) (table 1).

Of the available data on Pranayama practice among the symptomatic patients, 46.7% (28/60) practised it regularly. None of them required supplemental oxygen. Pranayama data on one of the eight patients who required supplemental oxygen was not available.

Table 2 outlines the various laboratory findings of the patients at admission. None of the patients had White blood cell count < 4 ×10^9^/L at admission. The platelet count in symptomatic patients was significantly higher than that of asymptomatic patients, although it was still within the normal range (table 2). Of the available data on platelet count among asymptomatic patients, 31.7% (13/41) had thrombocytopenia at presentation.

**Table 2.**
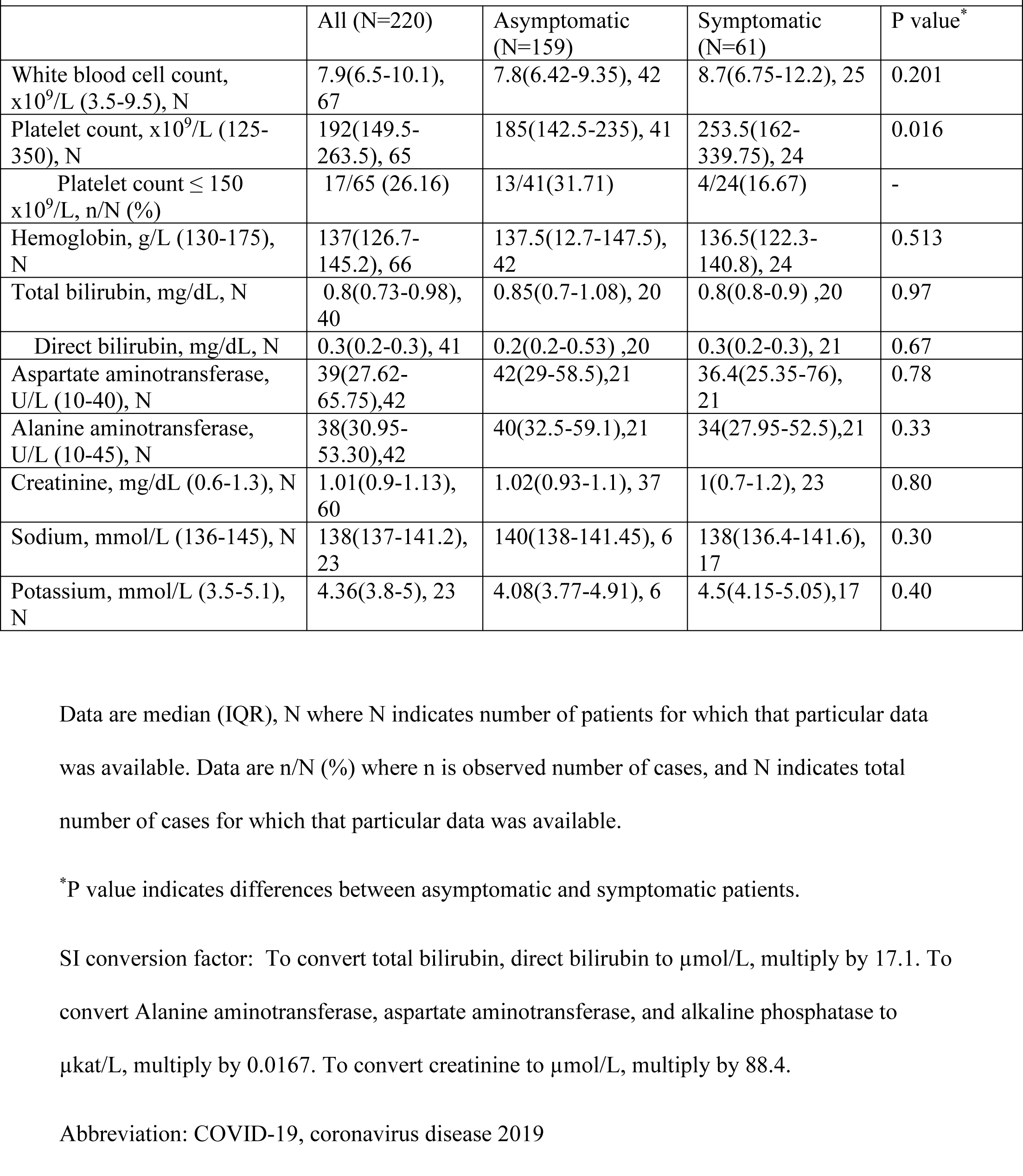
Laboratory findings of COVID-19 patients at admission, stratified by symptom status.

|  |  |  |  |  |
| --- | --- | --- | --- | --- |
|  | All (N=220) | Asymptomatic (N=159) | Symptomatic (N=61) | P value* |
| White blood cell count, $\times 10^9/L$ (3.5-9.5), N | 7.9(6.5-10.1), 67 | 7.8(6.42-9.35), 42 | 8.7(6.75-12.2), 25 | 0.201 |
| Platelet count, $\times 10^9/L$ (125-350), N | 192(149.5-263.5), 65 | 185(142.5-235), 41 | 253.5(162-339.75), 24 | 0.016 |
| Platelet count $\leq 150 \times 10^9/L$, n/N (%) | 17/65 (26.16) | 13/41(31.71) | 4/24(16.67) | - |
| Hemoglobin, g/L (130-175), N | 137(126.7-145.2), 66 | 137.5(12.7-147.5), 42 | 136.5(122.3-140.8), 24 | 0.513 |
| Total bilirubin, mg/dL, N | 0.8(0.73-0.98), 40 | 0.85(0.7-1.08), 20 | 0.8(0.8-0.9), 20 | 0.97 |
| Direct bilirubin, mg/dL, N | 0.3(0.2-0.3), 41 | 0.2(0.2-0.53), 20 | 0.3(0.2-0.3), 21 | 0.67 |
| Aspartate aminotransferase, U/L (10-40), N | 39(27.62-65.75), 42 | 42(29-58.5), 21 | 36.4(25.35-76), 21 | 0.78 |
| Alanine aminotransferase, U/L (10-45), N | 38(30.95-53.30), 42 | 40(32.5-59.1), 21 | 34(27.95-52.5), 21 | 0.33 |
| Creatinine, mg/dL (0.6-1.3), N | 1.01(0.9-1.13), 60 | 1.02(0.93-1.1), 37 | 1(0.7-1.2), 23 | 0.80 |
| Sodium, mmol/L (136-145), N | 138(137-141.2), 23 | 140(138-141.45), 6 | 138(136.4-141.6), 17 | 0.30 |
| Potassium, mmol/L (3.5-5.1), N | 4.36(3.8-5), 23 | 4.08(3.77-4.91), 6 | 4.5(4.15-5.05), 17 | 0.40 |

Data are median (IQR), N where N indicates number of patients for which that particular data was available. Data are n/N (%) where n is observed number of cases, and N indicates total number of cases for which that particular data was available.
\*P value indicates differences between asymptomatic and symptomatic patients.
SI conversion factor: To convert total bilirubin, direct bilirubin to $\mu\text{mol/L}$ , multiply by 17.1. To convert Alanine aminotransferase, aspartate aminotransferase, and alkaline phosphatase to $\mu\text{kat/L}$ , multiply by 0.0167. To convert creatinine to $\mu\text{mol/L}$ , multiply by 88.4.
Abbreviation: COVID-19, coronavirus disease 2019

Table 3 shows the treatment, complications and outcomes of the patients. 21.4% (47/220) received antibacterial drugs. None of the patients received any antiviral drugs or thromboprophylaxis. The median length of hospitalization was 14 days (IQR 13-14).

**Table 3.** Treatment, complications, and outcomes of COVID-19 inpatients stratified by symptom status.

|  |  |  |  |
| --- | --- | --- | --- |
|  | Total(N=220) | Asymptomatic (N=159) | Symptomatic (N=61) |
| **Treatment** |  |  |  |
| Antibiotics, n (%) | 47 (21.36) | 8 (5) | 39 (63.9) |
| Antiviral therapy, n (%) | 0 (0) | 0 (0) | 0 (0) |
| Thromboprophylaxis | 0 (0) | 0 (0) | 0 (0) |
| Hydroxychloroquine, n (%) | 1 (0.45) | 0 (0) | 1 (1.6) |
| Corticosteroids, n (%) | 9 (4.09) | 2 (1.3) | 7 (11.5) |
| Supplemental oxygen (including invasive mechanical ventilation), n (%) | 8 (3.63) | 0 (0) | 8 (13.1) |
| Needed invasive mechanical ventilation, n (%) | 3 (1.36) | 0 (0) | 3 (4.9) |
| Needed vasopressor, n (%) | 2 (0.9) | 0 (0) | 2 (3.3) |
| **Complications** |  |  |  |
| ARDS, n (%) | 3 (1.36) | 0 (0) | 3 (4.9) |
| Shock, n (%) | 2 (0.9) | 0 (0) | 2 (3.8) |
| Atrial fibrillation, n (%) | 1 (0.45) | 0 (0) | 1 (1.6) |
| **Admission to ICU, n (%)** | 8 (3.6) | 0 (0) | 8 (13.1) |
| **Median length of hospitalization in days (IQR)** | 14 (13-14) | 14 (13-14) | 14 (13-14) |
| **Outcomes** |  |  |  |
| Discharged to home after improvement, n (%) | 217 (98.63) | 159 (100) | 58 (95.1) |
| Referral to the other center, n (%) | 1 (0.45) | 0 (0) | 1 (1.6) |
| Death within hospital, n (%) | 2 (0.9) | 0 (0) | 2 (3.3) |

Data are n (%), where n is observed number of cases.
Column percentages might not round off to 100% due to rounding off.
Abbreviations: ARDS, acute respiratory distress syndrome; ICU, intensive care unit.

Of all patients, 3.6% (8/220) required supplemental oxygen of any kind via nasal canula, facemask, or mechanical ventilation. All of the patients who required supplemental oxygen were males. Among them, two patients needed vasopressors, with three requiring invasive mechanical ventilation. Two patients (0.9%) died.

## Discussion

In this first multicenter, prospective study on hospitalized adults with COVID-19 in Nepal, we found that most of the patients were asymptomatic, young males, migrant workers, with very few in-hospital deaths.

We observed that the males comprised an overwhelming majority (82.3%) of our patients This is similar to the data from other South Asian countries like India, Pakistan, Bangladesh, and Iran where the males comprised 60 to 90% of the admitted COVID-19 patients [9-12]. The male preponderance among the hospitalized patients was less pronounced in countries like the US, UK,China and Indonesia [13-16].One possible reason for this overwhelming male majority is that most of the COVID-19 patients in our study were migrant workers, and it is usually the adult males who go abroad for work. Another reason could be that the Nepalese society is highly patriarchal and are expected to earn a living for the family while most of the females are limited to their homes as housewives [17], thus limiting their exposure. Subgroup analysis of symptomatic patients showed that males were more likely to have severe disease and require supplemental oxygen than females. This is similar to the studies from abroad which showed severe disease and mortality is more common among males than females [15, 18, 19].

The patients in our study were relatively young with a median age of 31.5 years. This is similar to the data from other South Asian countries [10, 20, 21], but is in contrast to the studies from Italy, UK and USA [13, 14, 19]. This can partly be explained by the fact that the median age of the population in South Asian countries is substantially lower than countries like the US, UK or Italy [22].

We learnt that asymptomatic patients accounted for the vast majority (72.3%) of the study patients. Such high proportions of asymptomatic patients among hospitalized patients are similar to the studies from India [9, 23]. This may probably be because hospitals in Nepal do not have a uniform and well-followed criteria for admission and discharge. Resource limited countries have many unique challenges to deal with this pandemic [24]. Poor socio-economic conditions and overcrowding at home make it impossible for many patients to isolate themselves properly at home. There are inadequate isolation units at local places. All these reasons compelled us to admit even the asymptomatic patients, just for the sake of isolating them. In one case, we were forced to admit an asymptomatic patient because the neighbours and community leaders did not want anyone with COVID-19 living near them. This highlights the stigma associated with COVID-19 while also underscoring the limited knowledge about COVID-19 in the community.

We noted that low skilled migrant workers who had recently come from abroad represented nearly three-quarters of our patients. Most of them had returned from India. No other country in South Asia has reported such a big fraction of foreign migrant workers amongst hospitalized patients. This is reflective of the poor socio-economic situation in Nepal. One reason behind such predominance among foreign migrant workers could be that, until recently, the government had prioritized testing mainly of those persons who have recently returned from abroad and their contacts [5].

We spotted that compared to asymptomatic patients, symptomatic patients had a higher proportion of females (31.6% vs. 12.6%, p=0.001). This is in contrast to a study from China which showed that symptomatic patients had a higher proportion of males when compared to asymptomatic patients [25]. Other multiple studies from China and India showed no significant gender differences in relation to the symptom status [20, 26-28]. We believe our finding was due to the reporting bias and lower threshold of females to describe a particular sensation as being a symptom [29], as well as the fact that females are more likely to experience common somatic symptoms [30]. Further studies are needed to validate these differences.

We noticed that symptomatic patients had a higher median platelet count (253 ×10^9^/L vs. 185 ×10^9^/L, p=0.016) at presentation. However, the median platelet count was within the normal range in both groups. Studies from India and China did not find significant differences in platelet count in relation to symptom status [20, 26, 28]. Our finding is in contrast to the overall trend of decreasing platelet count with increasing disease severity, found in the studies from the US, China and Korea [31-34]. One study from Turkey showed that the platelet count was significantly low in COVID-19 PCR positive cases compared to PCR negative cases [35]. We also learnt that 31.7% (13/41) of the asymptomatic patients had thrombocytopenia at presentation. This is similar to a study from India where 36% of the asymptomatic patients had thrombocytopenia [20]. None of the patients who required oxygen supplementation or died had thrombocytopenia at presentation. In contrast, a meta-analysis which included nine different studies, showed that the platelet count was significantly lower in COVID-19 patients with the severe disease when compared to those with non-severe disease, although the included studies had high heterogeneity between them [33]. These observations in our study related to the platelet count need to be interpreted with caution because of caveat that only 65 patients out of 220 had their data on platelet count available, and these data are only from the time of presentation to the hospital. Further studies with more emphasis on laboratory parameters are needed to validate these findings from Nepal.

Among the symptomatic patients, regular practice of Pranayama was significantly associated with patients not needing supplemental oxygen. While we did not find any original study exploring Pranayama practice in relation to COVID-19 severity, Pranayama has been found to be beneficial in patients with respiratory illness [36]. Our finding needs to be interpreted with a caution that we did not take into account the potential confounding factors such as age, comorbidities etc. Further appropriately designed studies with more emphasis on this practice are needed.

We observed that, compared to asymptomatic patients, symptomatic patients included a greater proportion of patients who had recently come from abroad (31.9% vs. 15.78 %, p=0.024). One study from China, reported mixed findings where some of the symptoms were more common in imported cases compared to local cases whereas some other symptoms were less common [37]. We did not find any study from other South Asian countries looking at this point.

We observed that being illiterate (no formal schooling at all), use of alcohol, subjective feelings of being stigmatized by society were significantly associated with patients being symptomatic at presentation. We did not find any other study from South Asia investigating such variables in relation to COVID-19. Further studies need to be done to substantiate these findings.

Most of our patients (90.7%) were vaccinated with Bacillus Calmette-Guerin (BCG) at their birth. We did not find any significant association between BCG vaccination status and being symptomatic or the severity of disease. This is consistent with the data from other studies [38]. However, the proportion of COVID-19 patients vaccinated with BCG was much lower (49.2%) in a study from India [23].

This study has several limitations. We had a large amount of missing data for a prospective study. We could not perform many useful laboratory tests such as D-dimer, serum ferritin, IL-6, CT chest etc. Even for the basic lab tests we obtained, we could not obtain it on all patients and as frequently as it would have been in a country with better health care standards. We did not analyze the laboratory parameters serially. The admission and discharge criteria were not uniform among the hospitals in this study and the patients were not followed up after their discharge. The analysis of sub-groups was unadjusted for potential confounders. Many of the variables were self-reported by patients and we could not independently verify them. We did not have a mechanism to ensure the uniformity of diagnostic, and clinical management measures across the hospitals.

## Conclusion

In this first study of its kind from a resource limited South Asian nation, we found that most of the patients were asymptomatic, young male migrant workers with poor socio-economic conditions. Efforts need to be made to minimize the stigmatization of COVID-19 patients. Nepal needs to have uniform criteria for managing COVID-19 patients based on the evidence from the local context. The practice of Pranayama was associated with protection from severe disease, but more studies are needed to endorse this. Our unique observation of higher platelet count in symptomatic patients when compared to asymptomatic ones, needs to be substantiated by further research.

## Data Availability

Additional data is available by emailing the corresponding author at.

## Acknowledgements

We thank all the patients and their families involved in this study. We are thankful to Dr Basanta Chaudhary from Rapti Provincial Hospital for helping us in data collection. We extend our sincere thanks to all the medical staffs working together to fight against SARS-Cov-2. We are thankful to Ministry of Social Development, Sudurpaschim Province, Nepal for partially supporting us with grant (Grant number 332-2020).

## Author Contributions

AC, UNS, JJ contributed to the conception of this study. AC, NT and SBK helped in designing this study. AC, NT, PP, KK, PKS, KHA, SKJ, RJ, SB, UNS, APT contributed in acquisition of the data. AC, SRT, SS, HRP helped in data analysis and interpretation. AC wrote the original draft, PP and JJ contributed during revision. NT, UNS, SBK, PKS, KK helped in critically reviewing and editing the manuscript. AC and JJ worked to obtain the grant support. HRP, SS, SRT, KHA, APT, SKJ, SB, RJ helped with administrative, technical, and material support. All authors approved the final version of the manuscript. AC is the guarantor and corresponding author for this work, and attests that all listed authors meets authorship criteria and that no others meeting the criteria have been omitted.

